# Antibody persistence and neutralising activity in primary school students and staff: prospective active surveillance, June to December 2020, England

**DOI:** 10.1101/2021.07.14.21260502

**Authors:** Georgina Ireland, Anna Jeffery-Smith, Maria Zambon, Katja Hoschler, Ross Harris, John Poh, Frances Baawuah, Joanne Beckmann, Ifeanyichukwu O Okike, Shazaad Ahmad, Joanna Garstang, Andrew J Brent, Bernadette Brent, Felicity Aiano, Zahin Amin-Chowdhury, Louise Letley, Samuel E I Jones, Meaghan Kall, Monika Patel, Robin Gopal, Ray Borrow, Ezra Linley, Gayatri Amirthalingam, Kevin E Brown, Mary E Ramsay, Shamez N Ladhani

## Abstract

**Introduction:** SARS-CoV-2 serological studies have so far focused mainly on adults. Public Health England initiated prospective, longitudinal SARS-CoV-2 sero-surveillance in schools across England after the first national lockdown, which allowed comparison of child and adult responses to SARS-CoV-2 infection over time.

**Methods:** Staff and students had venepuncture for SARS-CoV-2 antibodies in school during June, July and December 2020. Blood samples were tested for nucleocapsid (Abbott) and receptor binding domain (RBD) antibodies (in-house assay), and student samples were additionally assessed for live virus neutralising activity.

**Results:** In June 2020, 1,344 staff and 835 students were tested. Overall, 11.5% (95% CI: 9.4-13.9) and 11.3% (95% CI: 9.2-13.6; p=0.88) of students had nucleoprotein and RBD antibodies, compared to 15.6% (95% CI: 13.7-17.6) and 15.3% (95% CI: 13.4-17.3; p=0.83) of staff. Live virus neutralising activity was detected in 79.8% (n=71/89) of nucleocapsid and 85.5% (71/83) of RBD antibody positive children. RBD antibodies correlated more strongly with neutralising antibodies (r^s^=0.7527; p<0.0001) than nucleocapsid antibodies (r^s^=0.3698; p<0.0001). A median of 24.4 weeks later, 58.2% (107/184) participants had nucleocapsid antibody seroreversion, compared to 20.9% (33/158) for RBD (p<0.001). Similar seroreversion rates were observed between staff and students for nucleocapsid (p=0.26) and RBD-antibodies (p=0.43). Nucleocapsid and RBD antibody quantitative results were significantly lower in staff compared to students (p=0.028 and <0.0001 respectively) at baseline, but not at 24 weeks (p=0.16 and p=0.37, respectively).

**Conclusion:** RBD antibodies correlated more strongly with live virus neutralising activity. Most seropositive students and staff retained RBD antibodies for >6 months after SARS-CoV-2 infection.

## Introduction

SARS-CoV-2 first emerged in December 2020 and spread rapidly across the globe, causing more than 100 million cases within 12 months, with more than 2 million deaths, mainly among the elderly.^1,2^ In order to limit the rapid spread of the virus, many countries implemented national lockdown measures including school closures which, in England, began on 20 March 2020. Children, however, have a lower risk of disease, hospitalisation or death due to COVID-19 compared to adults.^3^ Indeed, most children exposed to SARS-CoV-2 remain asymptomatic or develop mild, transient infection with non-specific symptoms, which may often not be attributed to the virus, raising concerns that they may unknowingly transmit the virus to others.^4^

Following partial reopening of UK schools after national lockdown in June 2020, there were concerns that the large number of children gathering in close confinement with limited social distancing would provide a hub for infection, leading to widespread transmission, not only among the children themselves, but also to school staff, household members and, potentially, the wider community.^5,6^ In order to investigate SARS-CoV-2 infection and transmission in educational settings, Public Health England (PHE) initiated prospective active surveillance in 131 primary schools (5-11 years) across England. Some schools implemented weekly swabbing for participating staff and students, while others involved swabbing with blood sampling to measure serum SARS-CoV-2 antibodies.^7^

Antibody testing provides a robust measure of prior exposure to SARS-CoV-2 in individuals with symptomatic, asymptomatic or mild, transient infection, making it a useful surveillance tool. Antibodies, and neutralising antibodies in particular,^8^ are also important markers of immunity against re-infection. Recent longitudinal adult studies, mainly in healthcare workers, have shown that SARS-CoV-2 antibodies following SARS-CoV-2 infection lasts for at least 6-8 months,^9^ and is associated with up 90% protection against re-infection.^10^ Antibody persistence after infection is, therefore, an important component in assessing duration of protection. This is particularly the case in children who can have multiple re-infections with endemic coronaviruses, at least in part due to antibody waning.^11,12^ Evidence to date has demonstrated antibody persistence in children up to 62 days after SARS-CoV-2 infection.^13^ Here, we report the duration of SARS-CoV-2 antibodies against nucleocapsid (N) and the receptor binding domain (RBD) of the spike protein at least 6 months after infection in primary school students and staff taking part in the COVID-19 surveillance in school KIDs (sKIDs).^7^ Live virus neutralising activity was assessed in a subset of seropositive students and assessed for correlation with N and RBD antibodies.

## Methodology

### Sampling

The protocol for sKIDs surveillance is available online.^7^ Briefly, 131 primary schools were recruited and included 46 schools involved in swabbing with blood sampling at three time-points: following partial reopening of schools on 01 June 2020 (1-19 June 2020) and end of the summer term (3-23 July 2020), which were used as baseline seroprevalence and, following full reopening of all schools on 1 September 2020, at the end of the autumn term (23 November to 18 December 2020).

### Laboratory testing

Blood samples were tested for nucleoprotein antibodies using a chemiluminescent microparticle immunoglobulin G (IgG) immunoassay targeting the nucleocapsid (SARS-CoV-2 IgG, Abbott Commerce Chicago, USA) with a seropositivity threshold of 0.8. The samples were also tested for RBD antibodies using an in-house indirect IgG RBD assay.^14^ Blood samples with sufficient serum from N antibody positive students in June and July 2020 were also tested for neutralising antibodies using a modification of the WHO influenza microneutralisation methodology.^15^

### Analysis

Data were analysed using Stata SE (version 15.1). Seropositivity for N and RBD antibodies was analysed using an index value for each test. Antibody persistence at 24 weeks was estimated for participants tested in June 2020 and at the end of the autumn term in November/December 2020. Where participants were also tested in July, antibody persistence at 4 weeks was estimated. Seroreversion was defined as testing negative after previously testing positive on the same antibody assay platform. Categorical variables were described as proportions and compared using Chi^2^ and Fisher’s Exact tests. Spearman’s rank was used to test for correlation between neutralising antibody response, N-antibody and RBD-antibody results. Correlation between longitudinal samples, taken on average 4 weeks later, was also tested for. Kruskal-Wallis H Test was used to compare neutralising results by age-group in children. Seroprevalence and antibody persistence proportions were calculated with 95% binomial confidence intervals. Quantitative antibody results were log-transformed and compared using matched and unmatched t-tests. A linear regression on log-transformed quantitative results was used to test for differences between students and staff, and at baseline and final sample, including an interaction between participant type and testing round. For RBD, this only included participants who were RBD positive at baseline. Samples were clustered by individual participant to account for repeated measurements. The coefficient was exponentiated to provide ratio measures.

### Ethics approval

The protocol was approved by PHE Research Ethics Governance Group (Ref: NR0209, 16 May 2020)

## Results

### Seroprevalence in June

At recruitment in June 2020, 2,179 samples obtained from sKIDs participants in England had sufficient sample volume to be additionally tested for RBD antibodies (Figure 1). Staff represented 61.7% (1,344/2,179); median age was 43 years (IQR, 33-52) and 80.8% (1,086/1344) were females. The median age of 835 students (38.3%) was 8 years (IQR: 6-10) and 50.5% (422/835) were females.

**Figure 1:**
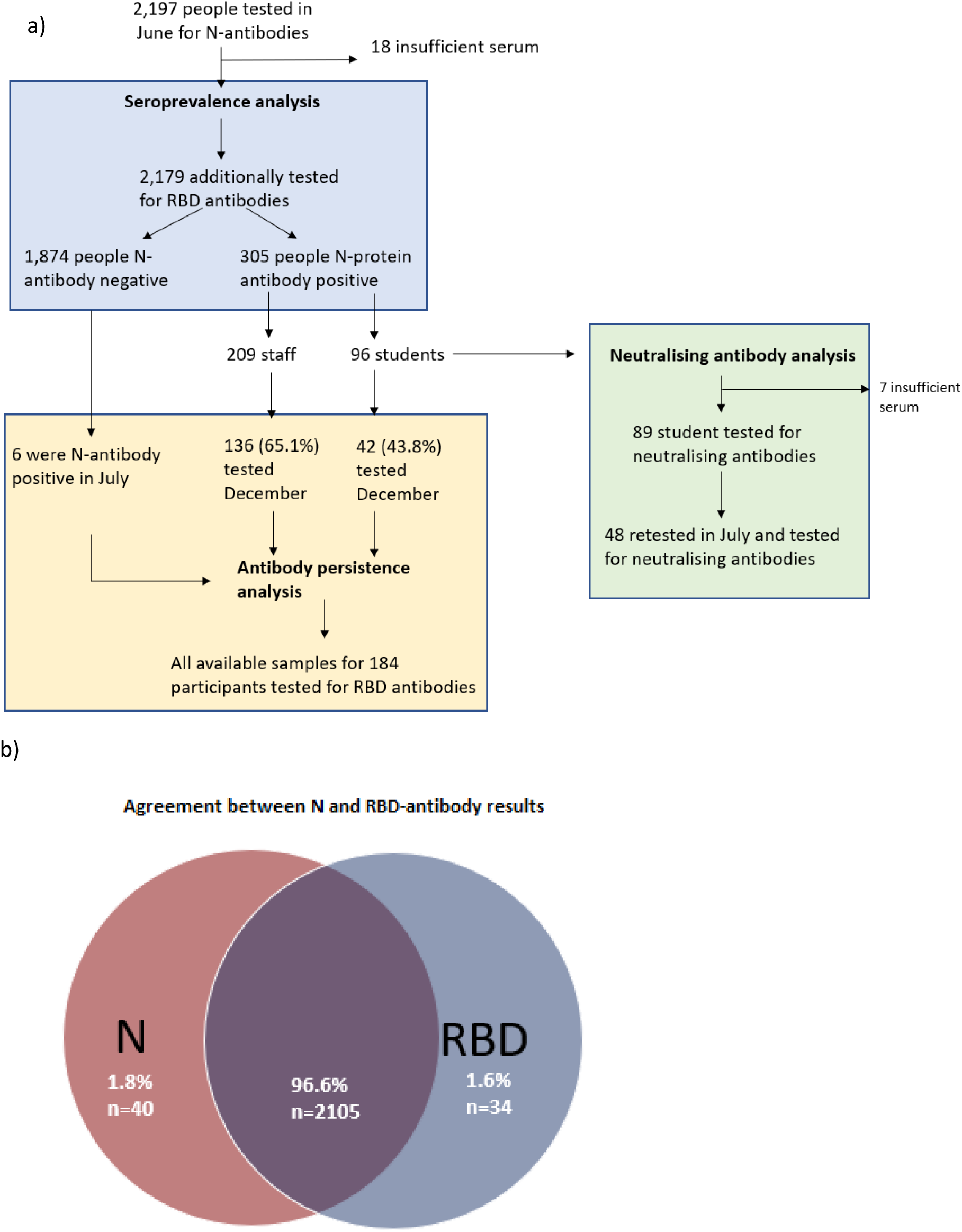
Participant flow between antibody seroprevalence, antibody persistence and neutralising antibody analysis in sKIDs participants (a) and agreement between N and RBD antibody results (b).

Overall, seropositivity for N (14.0%; 305/2179; 95% CI: 12.6-15.5) and RBD (13.7%; 299/2179; 95% CI: 12.3-15.2) antibodies was similar (p=0.79). This was also true for students and staff: 11.5% (95% CI: 9.4-13.9) and 11.3% (95% CI: 9.2-13.6; p=0.88) of students were seropositive for N and RBD antibodies, respectively, compared to 15.6% (95% CI: 13.7-17.6) and 15.3% (95% CI: 13.4-17.3; p=0.83), respectively, among staff (**Table 1**). N and RBD-antibody quantitative results correlated significantly (0.4280, p<0.0001) (Figure 2).

**Table 1:** Nucleocapsid and RBD antibody seropositivity at baseline in primary school students and staff during June 2020.

|  |  | RBD antibody |  |  |  |  |  |  |  |
| --- | --- | --- | --- | --- | --- | --- | --- | --- | --- |
|  |  | Total |  |  | Students |  |  | Staff |  |
|  |  | Negative | Positive |  | Negative | Positive |  | Negative | Positive |
| Nucleocapsid antibody | Negative | 1840 (84.4) | 34 (1.6) |  | 733 (87.8) | 6 (0.7) |  | 1107 (82.4) | 28 (2.1) |
|  | Positive | 40 (1.8) | 265 (12.2) |  | 8 (1.0) | 88 (10.5) |  | 32 (2.4) | 177 (13.2) |

**Figure 2:**
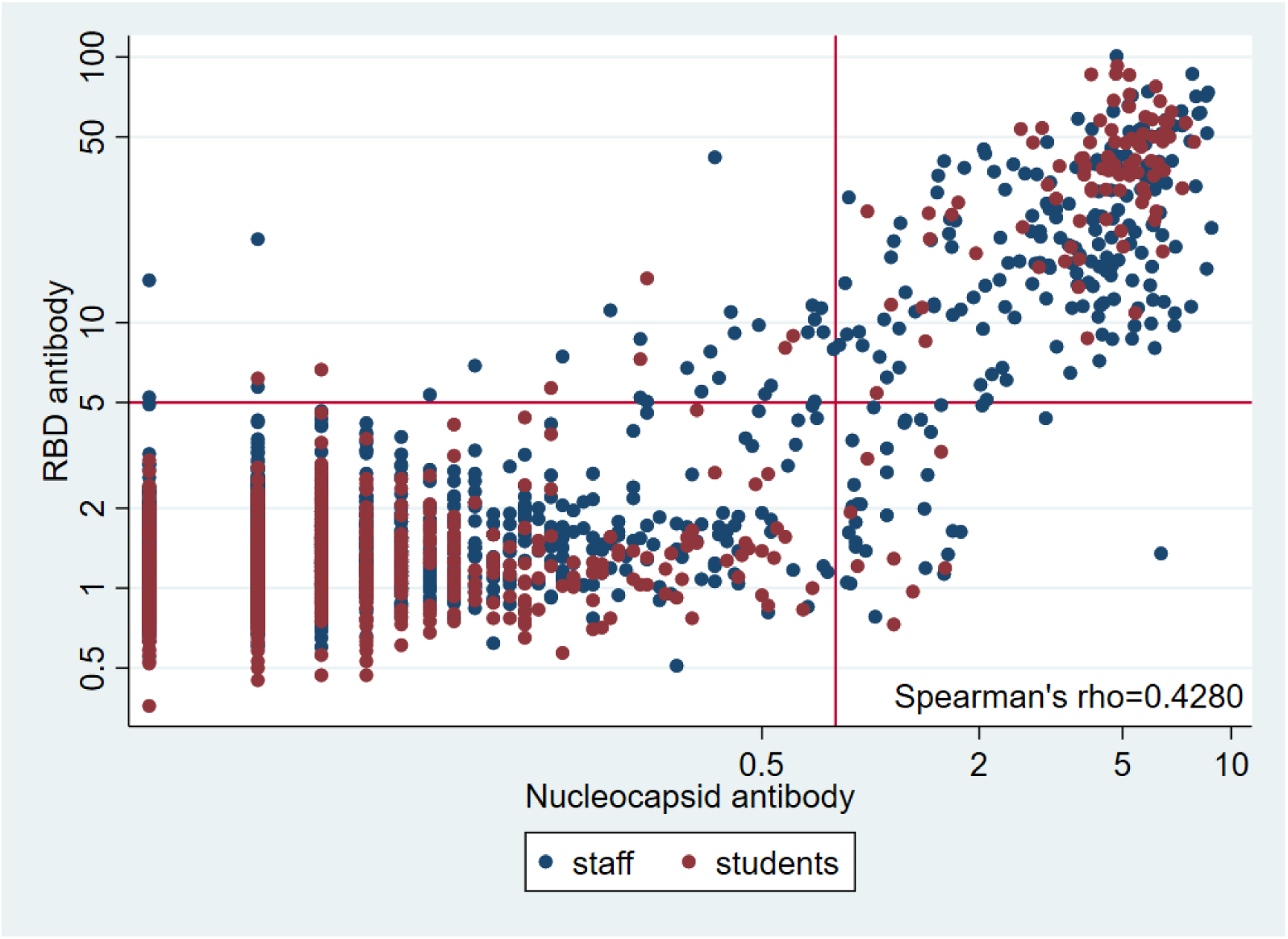
Log nucleocapsid and RBD antibody titres for 2,179 students and staff tested as part of the sKIDs study in June 2020 in England. Lines denote the threshold values for reporting a positive result. Neutralising antibodies in students: correlation with Nucleocapsid and RBD antibodies

N and RBD-antibody result agreement was 96.6% (Figure 1b), with a lower proportion of staff having concordant results compared to students (95.5% vs 98.3%; p<0.001). Of the 74 (3.4%) with discordant results, 34 (45.9%) were N-antibody negative but RBD-antibody positive, 40 (54.1%) were N-antibody positive but RBD negative and there was no difference in proportion with discordant results between students and staff (p=0.52).

Live virus neutralising antibody titres were assessed in 89 N-antibody positive students from the June testing round. In these samples, 79.8% (71/89) had neutralising antibodies and all six children (6.7%) who were RBD antibody negative at baseline had no detectable neutralising antibodies. Of the 83 RBD-antibody positive students at baseline, 85.5% (71/83) had neutralising antibodies. One month later, a paired sample was available and tested for neutralising activity in 48 students. Across all samples (n=142), both N and RBD quantitative results positively correlated with neutralising titres, with RBD antibodies demonstrating a stronger correlation (N: 0.3698, p<0.0001 and RBD: 0.7527, p<0.0001). (**Supplementary Figure 1a&1b**). There was no difference in neutralising antibody titres by age in students (p=0.52) (**Supplementary Figure 2**).

Median neutralising antibody titres in 48 students with paired samples in June and July were similar (35.2 [IQR,26.0-54.1] in June vs. 33.0 [IQR:22.5-49.8] in July; p=0.23) and correlated strongly between the two-time points (0.8087 p<0.0001) (**Supplementary Figure 1c**).

### Nucleocapsid and RBD-antibody persistence

Of the 311 participants (211 staff, 100 students) who were N-antibody positive in June 2020 or seroconverted by July 2020, 184 (59.2%) underwent subsequent sampling at the end of the autumn term. This cohort included 178 participants who were N-antibody positive in June and 6 who had seroconverted by July (Figure 1).

Of the 179 who were N-antibody positive in June, 69.7% (124/178) had a further antibody sample in July (median 4.3 weeks, IQR: 3.9-5.1). Of these, none of the students seroreverted for N (0/25) or RBD (0/24) antibodies, whereas, 5.1% (5/99) of staff seroreverted for N and 10.0% (8/80) for RBD antibody (Figure 2). Only one of the 5 staff participants with N antibody seroreversion remained positive for RBD antibodies.

At the end of the autumn term, 184 (46 students, 138 staff) had a repeat blood sample at a median of 24.4 weeks (IQR: 23.1-25.9) later. Of these, 58.2% (107/184) seroreverted for N-antibodies compared to 20.9% (33/158) for RBD (p<0.001) (**Figure 3**). There was no difference in N (65.2% vs 55.8%, p=0.26) and RBD antibody (16.7% vs 22.4% p=0.43) seroreversion between students and staff. Of those who seroreverted on the N antibody assay, 73.3% (n=22/30) of students and 59.7% (n=46/77; p=0.19) of staff remained positive for RBD antibodies.

**Figure 3:**
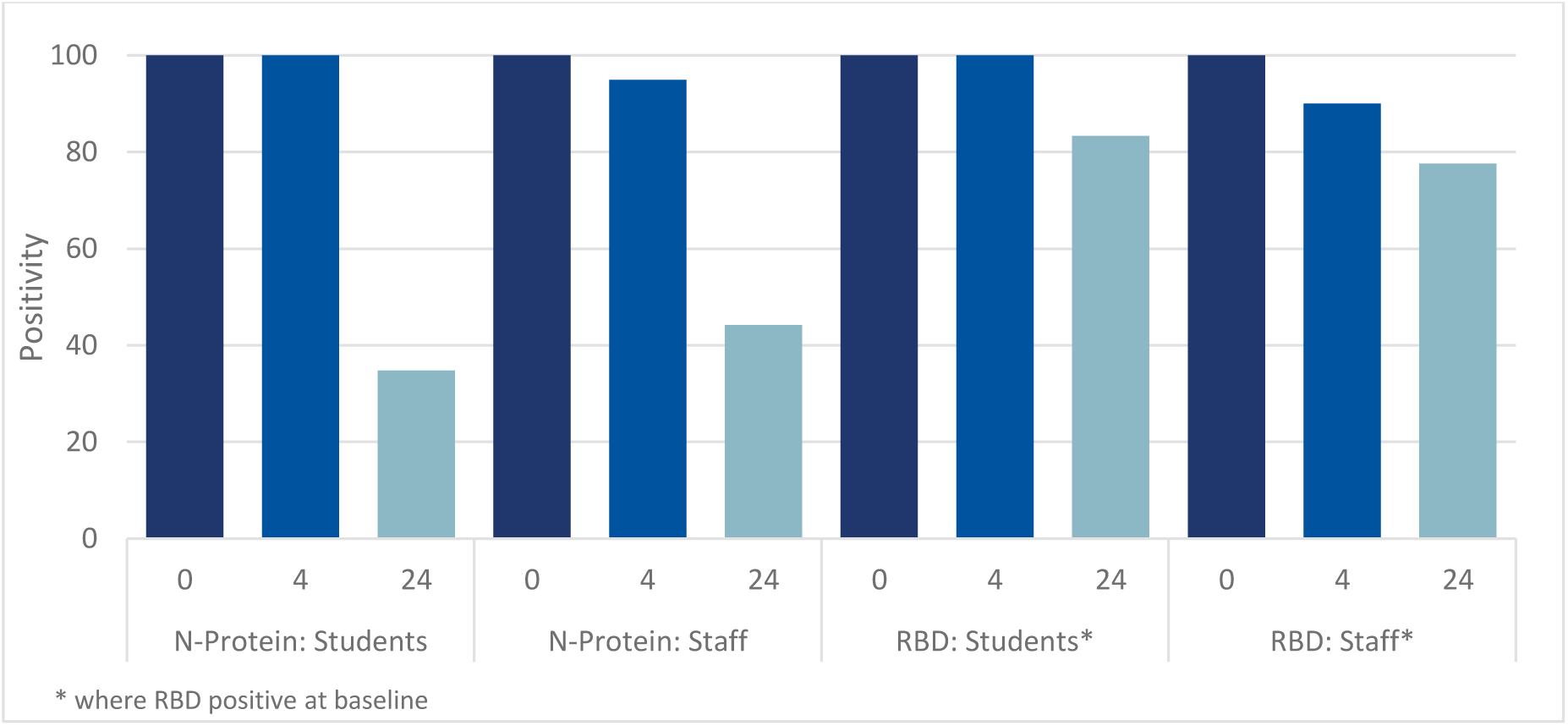
Nucleocapsid and RBD-antibody^*^ positivity at 0, 4 and 24 weeks in the sKIDs Study in England. ^*^where RBD positive at baseline

In this cohort of 184 N antibody positive participants, 26 (14.1%) had a negative RBD result in June 2020, including 4 (4/46, 8.7%) students and 22 (22/138, 15.9%) staff (p=0.22). Only one, a staff member (1/22, 4.5%), subsequently seroconverted on the RBD assay by the end of the autumn term.

The N and RBD antibody quantitative results were significantly lower in staff compared to students (p=0.028 and <0.0001 respectively) at baseline, but not at 24 weeks (p=0.16 and p=0.37, respectively) (Figure 4). Using linear regression, N-antibody results at baseline were 22.8% (95% CI: 6.0-35.0%) lower in staff than students and 42.1% (95% CI: 29.3-52.6) lower against RBD. After 6 months, N-antibody results were 88.2% (95% CI: 83.4-91.7) lower compared to baseline results in students and 78.4% (95% CI: 75.0-81.4) lower among staff. The corresponding reductions for RBD antibody results were 69.0% (95% CI: 58.5-76.9) and 50.4% (95% CI: 43.7-56.4), respectively. Only 3.3% (6/184) and 10.8% (17/158) had an increase in N and RBD quantitative results respectively at 24 weeks than baseline.

**Figure 4:**
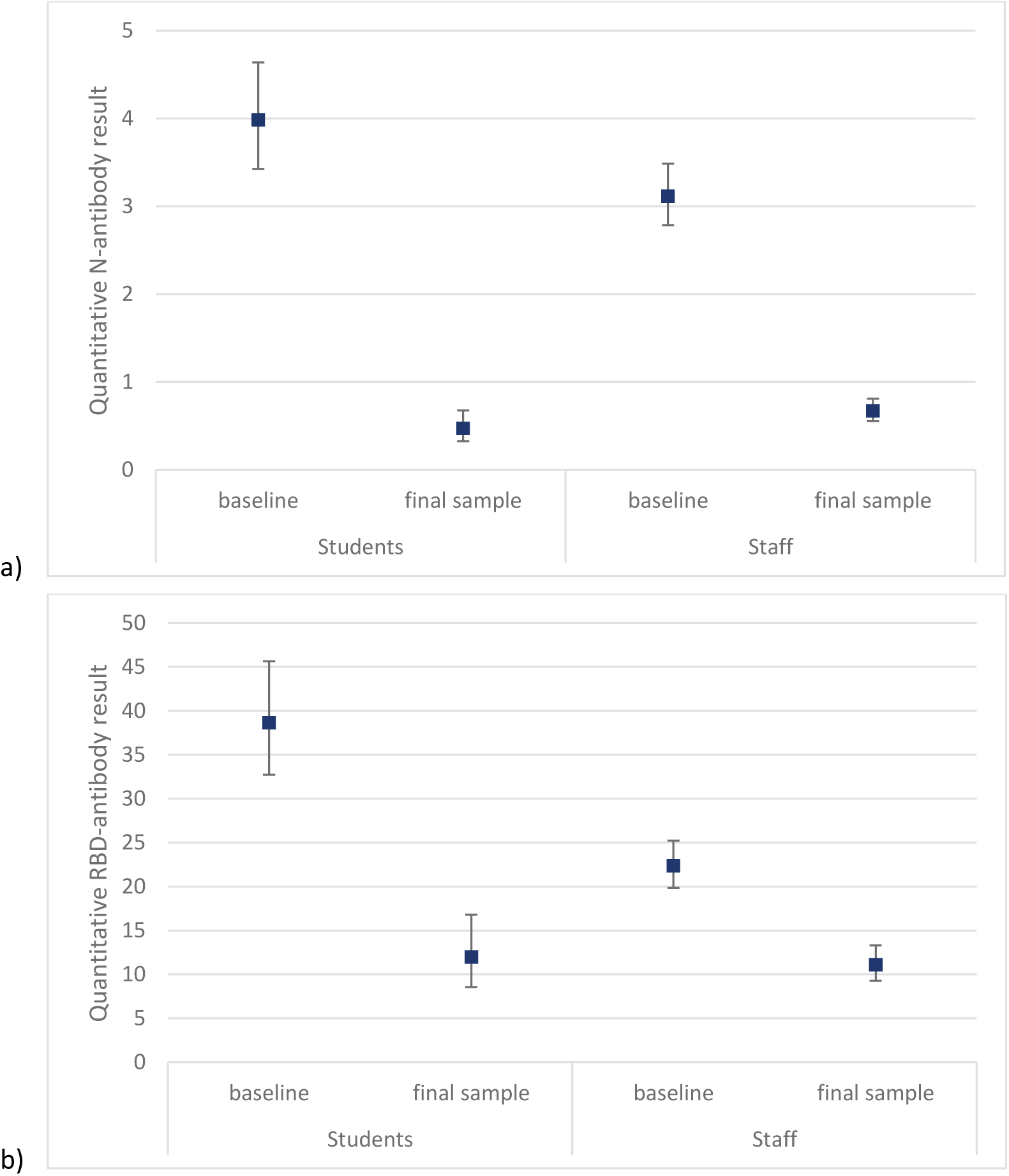
Geometric mean, with 95% confidence intervals, of (a) nucleocapsid and (b) RBD^*^ antibody titres at baseline and 24 weeks (final sample) for students and staff participating in the sKIDs study. ^*^where RBD antibody positive at baseline

## Discussion

SARS-CoV-2 antibody prevalence was similar in primary school staff and students in June 2020, with high concordance between antibody seropositivity against N and the RBD of the spike protein. Antibody kinetics over time, however, differed. Less than half of positive participants retained N-antibodies measured using the Abbott assay. RBD-antibodies, on the other hand, correlated more strongly with neutralising antibody titres, and persisted in 78% of students and 83% of staff for more than 6 months after infection. Antibody results at baseline were significantly higher in students than staff but declined over time such that they were similar in the two groups after 24 weeks.

The Abbott N-antibody assay was originally selected for SARS-CoV-2 surveillance in educational settings because it could detect SARS-CoV-2 antibodies within 7-14 days after infection, earlier than assays measuring Spike protein antibodies, thus facilitating early detection of seroconversions after symptomatic or asymptomatic infections between testing rounds.^6,16,17^ There is, however, increasing evidence that the high seroreversion rates observed with the Abbott assay are not biological but a function of the antibody assay. In our other adult cohort studies, for example, 44% of adults seroreverted for N-antibodies using the Abbott assay by 9 weeks and 58% by 24 weeks,^16^ but N-antibodies remained positive using other assays such as the Roche N assay.^16^ Other investigators have also reported similar findings.^18^ Nucleocapsid is an immunodominant antigen in many enveloped virus infections and, although non-neutralising, nucleoprotein antibodies to other viruses have been shown to facilitate viral clearance *in vivo*, most likely through T-cell mediated immunity and cytotoxic T cells (CTLs) against nucleoprotein, which can rapidly eliminate infected cells displaying nucleoprotein peptides.^19^

Because of the high seroreversion rate for N-antibodies with the Abbott assay, we re-tested all available sera with a validated in-house RBD assay, which we and others have shown to correlate more strongly with neutralising antibodies in adults.^14,20,21^ We have now also confirmed this strong correlation in young children. Moreover, other investigators found almost all adults with confirmed COVID-19 had neutralising antibodies up to 6 months later and, while antibody results decreased progressively over time, anti-RBD antibody concentrations and neutralising antibody titres remained strongly correlated at 1, 3 and 6 months after infection.^22^ This is reassuring for our cohort as the majority of students and staff will have been infected with SARS-CoV-2 during the first pandemic peak in the UK in March and April 2020, and retained RBD antibodies up to nine months later, at the end of the autumn term in November/December 2020.

A small proportion of participants in our cohort had detectable N-antibodies but were RBD antibody negative upon initial testing and, importantly, had no virus neutralising activity and remained RBD-antibody negative in subsequent tests. Taken together, these findings suggest that a small proportion had false positive antibody results using the Abbott assay, potentially due to non-specific, cross-reactive antibodies from prior coronavirus infections.^16^The choice of antibody assay in sero-surveillance will become increasingly important as current vaccines only induce spike protein antibodies, while measurement of N-antibodies will help confirm prior SARS-CoV-2 infection irrespective of vaccination status.

Compared to staff, students had significantly higher RBD and N antibody results at baseline, but these differences had resolved after six months, with similar RBD antibody results and seroreversion rates during this period. This is reassuring, given that 76% of student compared to 47% of staff who seroconverted during the surveillance period had asymptomatic infection, indicating that children with asymptomatic SARS-CoV-2 infection are able to mount a robust and durable immune response similar to – if not better than – adults.^7^

We assessed the association between antibody results and virus neutralising activity in students only because this has already been extensively investigated in adults. The few studies in children have reported robust immune responses following SARS-CoV-2 infection *albeit* with some important differences when compared with adults.^23,24^ Among 1-24 year-olds in a recent New York seroprevalence study, for example, SARS-CoV-2 total IgG and RBD antibody titres were highest in younger children and declined with increasing age, while surrogate neutralising antibody activity and antibody avidity were lowest in younger children and increased with age.^24^ Others have also reported lower neutralising antibody titres in children compared to adults.^23–25^ We have, however, recently reported a higher prevalence and magnitude of cellular responses against the spike protein of SARS-CoV-2 in in our cohort of primary school-aged children compared to adults more than six months after primary infection.^26^

### Strengths and limitations

The early initiation of surveillance in primary schools provided a unique opportunity to monitor seroprevalence, seroconversion and antibody persistence in more than 2,000 healthy young children and adults with similar exposure risks to SARS-CoV-2 in 45 schools across England.^7^ A limitation of our study was the limited testing for SARS-CoV-2 infection in the community during the first wave of the pandemic; we were, therefore, unable to confirm acute SARS-CoV-2 infection in symptomatic participants prior to recruitment. Additionally, most seropositive children in June 2020 were reported by their parents to be asymptomatic and, therefore, the timing of their infection was not known. Additionally, we only assessed the correlation of N and RBD antibodies with neutralising activity during the first two rounds of testing in June and July 2020 and assumed that this correlation would be retained in round 3 in December 2020, as has been reported by others. Moreover, whilst we focused on the antibody responses to infection in this analysis, cellular immune responses are also likely to play an important role in protection against SARS-CoV-2 re-infection.^26^ Finally, this surveillance was undertaken prior to the emergence and rapid spread of the alpha and delta SARS-CoV-2 variants, which have both been associated with increased transmission compared to previously circulating strains.^27,28^ We, therefore, cannot comment on the protective effects of prior SARS-CoV-2 infection against reinfection with new variants.

## Conclusion

The majority of primary school students and staff retained RBD antibodies, which strongly correlated with neutralising activity, for more than 6 months after SARS-CoV-2 infection. Our findings provide further evidence of a robust and sustained immune response in children following primary SARS-CoV-2 infection. Further studies are needed to assess protection against emerging variants of concern.

## Data Availability

Applications for relevant anonymised data should be submitted to the Public Health England Office for Data Release

https://www.gov.uk/government/publications/accessing-public-health-england-data/about-the-phe-odr-and-accessing-data.

## Acknowledgements

The authors would like to thank the schools, headteachers, staff, families and their very brave children who took part in the sKIDs surveillance. The authors would also like to thank Sir Jeremy Farrar, Sir Patrick Vallance, members of the Department for Education, Department of Health and Social Care, London School of Hygiene and Tropical medicine (LSHTM), Office for National Statistics (ONS) and the UK Scientific Advisory Group for Emergencies (SAGE) for their input and support for the sKIDs surveillance.

## Author Contributions

SNL, FB, JB, IO, SA, JG, AB, BB, GA, VS, JLB, KB and MR were responsible for conceptualization and study design/methodology. SNL, FB, JB, IO, SA, JG, AB, BB, GI, FA, ZAC, LL, JF, SEIJ, RB, EL, MZ, AJS, KH, JP contributed to project administration (including laboratory colleagues). SNL, AJS, MZ and GI contributed to the original draft and GI and SNL conducted the formal analysis and were responsible for data validation. All authors contributed to reviewing and editing the manuscripts.

## Data sharing

*Applications for relevant anonymised data should be submitted to the Public Health England Office for Data Release: https://www.gov.uk/government/publications/accessing-public-health-england-data/about-the-phe-odr-and-accessing-data*.

