## Supplementary Materials for "Antibody persistence and neutralising activity in primary school students and staff: prospective active surveillance, June to December 2020, England"

### Supplementary Figures

*Supplementary Figure 1: Correlation between neutralising antibody quantitative result and N-protein (a) and RBD (b) antibody quantitative results and neutralising antibody quantitative results in June (time 1) and July (time 2) (c). Red lines denote the threshold values for reporting a positive result and brown line is where x=y.*

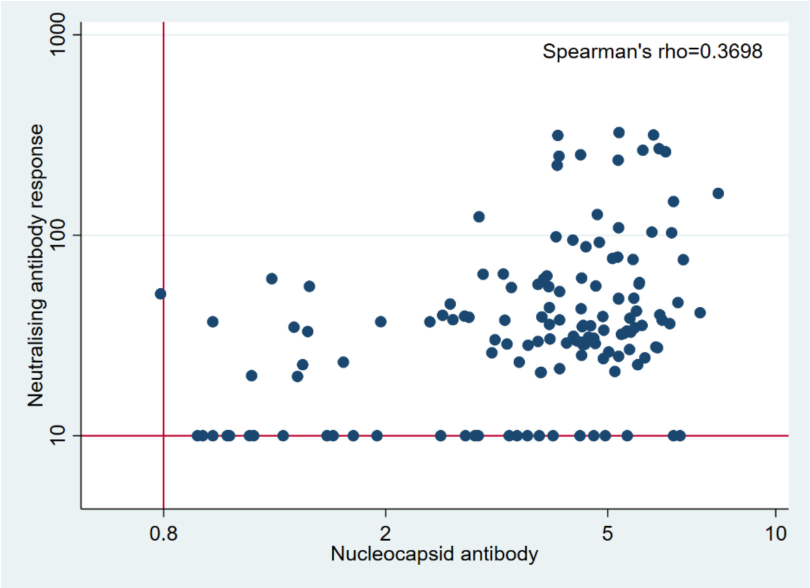


*a)*

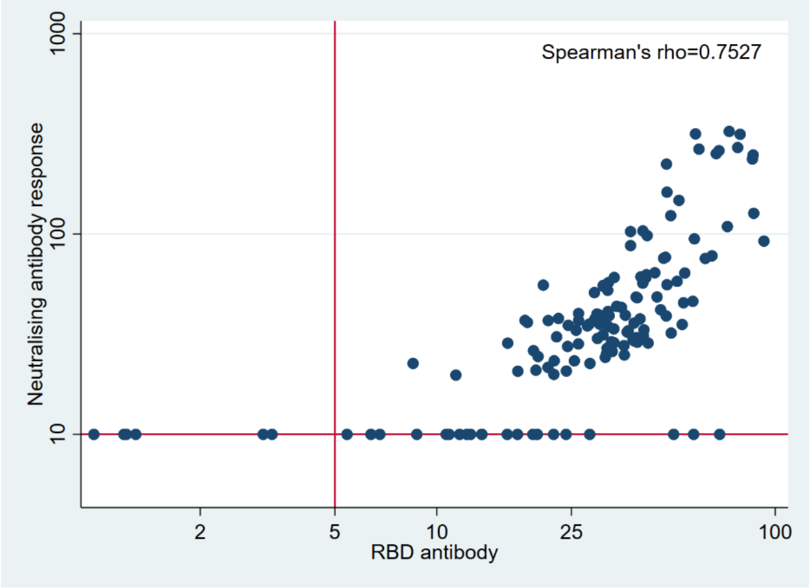


b)

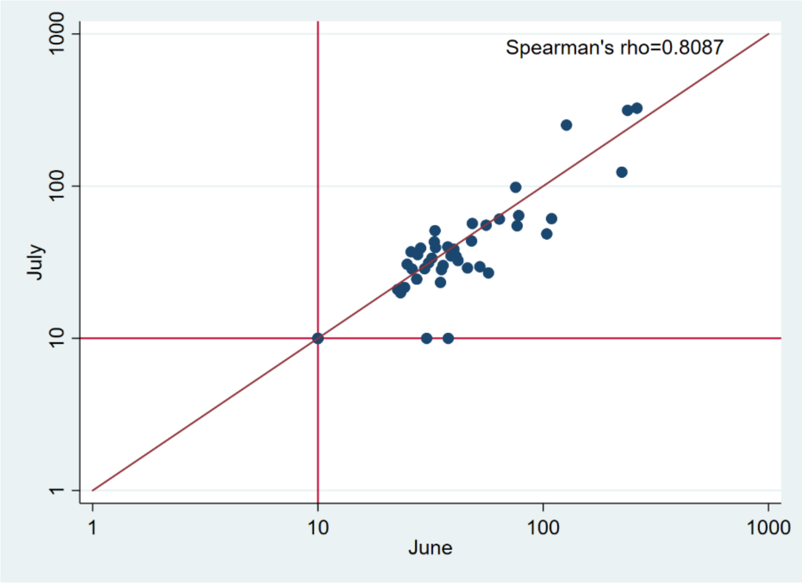


c)

*Supplementary Figure 2: Violin Plot of neutralising antibody quantitative result by age (in years) on first N-protein positive sample (n=89)*

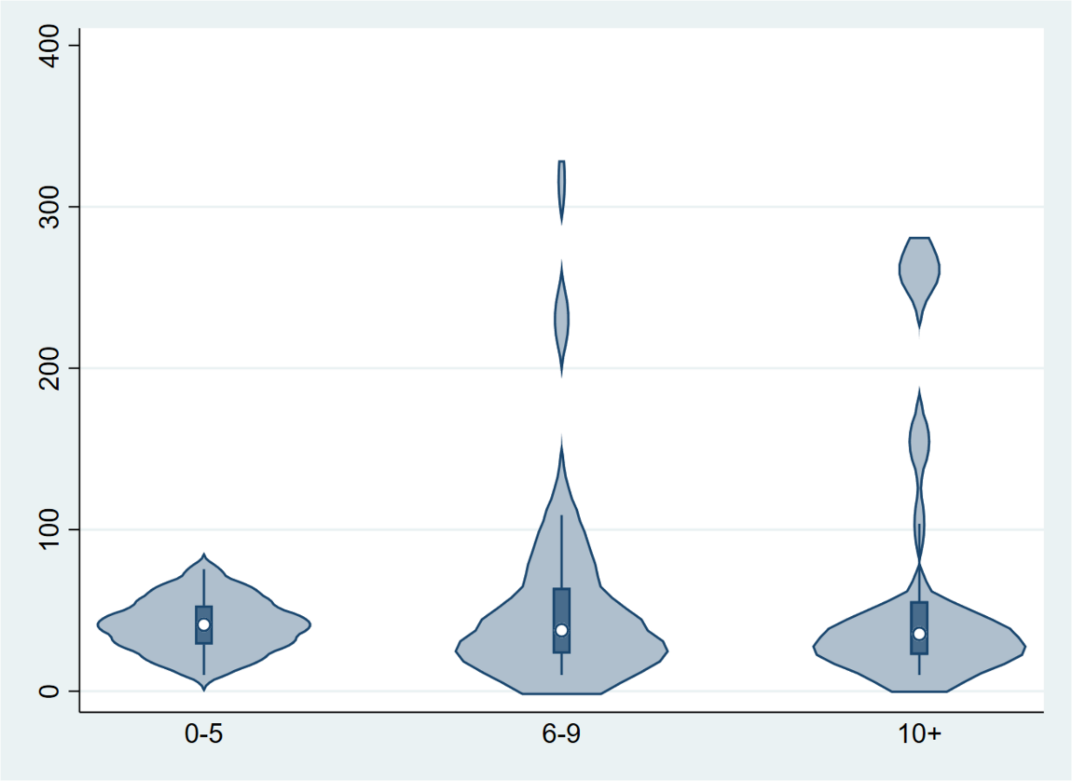
